# Phenotype and genetic analysis of data collected within the first year of NeuroDev

**DOI:** 10.1101/2022.08.22.22278891

**Authors:** Patricia Kipkemoi, Heesu Ally Kim, Bjorn Christ, Emily O’Heir, Jake Allen, Christina Austin-Tse, Samantha Baxter, Harrison Brand, Sam Bryant, Nick Buser, Victoria de Menil, Emma Eastman, Serini Murugasen, Alice Galvin, Martha Kombe, Alfred Ngombo, Beatrice Mkubwa, Paul Mwangi, Collins Kipkoech, Alysia Lovgren, Daniel G. MacArthur, Brigitte Melly, Katini Mwangasha, Alicia Martin, Lethukuthula L. Nkambule, Alba Sanchis-Juan, Moriel Singer-Berk, Michael E. Talkowski, Grace VanNoy, Celia van der Merwe, The NeuroDev Project, Charles Newton, Anne O’Donnell-Luria, Amina Abubakar, Kirsten A Donald, Elise Robinson

## Abstract

**Summary:** Genetic association studies have made significant contributions to our understanding of the aetiology of neurodevelopmental disorders (NDDs). However, the vast majority of these studies have focused on populations of European ancestry, and few include individuals from the African continent. The NeuroDev project aims to address this diversity gap through detailed phenotypic and genetic characterization of children with NDDs from Kenya and South Africa. Here we present results from NeuroDev’s first year of data collection, including phenotype data from 206 cases and clinical genetic analysis of 99 parent-child trios. The majority of the cases met criteria for global developmental delay/intellectual disability (GDD/ID, 80.3%). Approximately half of the children with GDD/ID also met criteria for autism, and 14.6% met criteria for autism alone. Analysis of exome sequencing data identified a pathogenic or likely pathogenic variant in 13 (17%) of the 75 cases from South Africa and 9 (38%) of the 24 cases from Kenya, as well as 7 total cases with suspicious variants of uncertain significance (VUS) in emerging disease genes that were matched through the MatchMaker Exchange. Data from the trio pilot cases has already been made publicly available, and the NeuroDev project will continue to develop resources for the global genetics community.

## Introduction

The Genetic Characterization of Neurodevelopmental Disorders project (NeuroDev) is a study of neurodevelopmental disorders (NDDs) that will collect and analyze extensive genetic and phenotypic data from over 5000 people, including ∼3600 children and their parents, in Kenya and South Africa over the next several years.^1^ Based on existing recruitment patterns, most of the 2000 case individuals enrolled in the study are projected to meet criteria for global developmental delay/intellectual disability (GDD/ID) or autism. All data (e.g., phenotypes; genotype array and exome sequencing data) and materials (e.g., blood DNA; cryopreserved cell lines or CPLs) generated by NeuroDev will be made publicly available through approved National Institute of Mental Health repositories. Through the data collection activity, which includes African ancestry populations largely absent from genetic reference panels (e.g., gnomAD), and through the public release of deeply characterized case and control data, NeuroDev aims to support diversity in biomedical research.

In this paper, we present the NeuroDev Pilot, which includes data from the project’s first collection year. During the first year, we collected data from more than 200 cases and 600 total participants. We describe in this report phenotype data from all cases collected in the pilot period, along with genetic analysis of 99 exome-sequenced trios. Supplementing these early genetic and phenotypic findings, we present learning points of the first year of data collection and modifications made to the NeuroDev protocol initially presented in de Menil et al.^1^ We hope these reflections on process will help others in the design and execution of similar projects, as more work on NDDs in Africa is both needed and underway. At only 99 trios, the NeuroDev trio pilot data is now the largest African NDD collection for which genetic and phenotypic data are publicly available to the research community. The trio data presented here can be accessed through National Human Genome Research Institute (NHGRI) Analysis Visualization and Informatics Lab-space (ANVIL) controlled access data repository (https://anvilproject.org/data/).

## Results

### Data Collection

From August 2018 to July 2019, we enrolled 219 cases, 195 case mothers, 115 case fathers, and 92 unrelated child controls. There were 106 total parent-case trios, and an additional 113 cases had only one participating parent. In Kilifi County, Kenya, case families were recruited from specialized neurology and occupational therapy clinics, special needs schools and from a database of previous studies. The case children enrolled in the study had tentative NDD diagnoses obtained from checklists or previous neuropsychological assessments. These diagnoses were confirmed by a study clinician prior to enrollment. In Cape Town, South Africa, participants with a clinical NDD diagnosis were recruited from the developmental clinic at Red Cross Children’s Hospital and Tygerberg Hospital. Cases included in the study had a diagnosis of any NDD (excluding primary motor disorders, e.g. cerebral palsy) and were between 2 and 18 years of age. We present in this report phenotype analysis of all 219 cases, along with genetic data analysis of the first 100 trios, 99 of which passed quality control measures for analysis.

At the end of the project’s first year, both the trio collection rate and the overall case collection rate in NeuroDev were aligned with our four-year sample size targets, and phenotype battery completion was high. In the first year, all participants had data for the demographic, neuromedical assessment, and behavioral measures. The behavioral measures included the Social Communication Disorders Checklist (SCDC)^2^ ; 3Di Brief^3^ to measure of autism characteristics; and Swanson, Nolan, and Pelham Rating Scale (SNAP-IV) to measure canonical ADHD symptomatology.^4^ Item level missingness was below 10% for all measures (for details, see Supplementary Table 1 and 2).

The Raven’s Progressive Matrices (RPM), NeuroDev’s nonverbal reasoning ability measure, was completed by 99% of participating parents. Inspired by the Simons Simplex Collection (SSC) cognitive testing approach,^5,6^ all enrolled children aged 6 years and older were offered the opportunity to attempt the RPM^7^ (Standard RPM if 12 years or older; Colored RPM if 6-11 years), which was adapted and validated for use in Kilifi.

Many case children (49%) could not complete age-appropriate versions of the RPM due to the extent of their developmental delay or behavioral challenges. All case children under 6 years of age and case children over 6 years who could not complete the RPM were offered the Molteno Adapted Scales of Development (Molteno). We experienced similar challenges in some cases with regards to the completion of the Molteno, though all participants have at least some data. This experience is common to studies of ID and autism, particularly those that include children and severe phenotypes.^8,9^ Missing values from the RPM are associated with case severity and will therefore be informative.

In the first year of data collection, trio family ascertainment was higher than anticipated. The goal in South Africa to recruit 100 trios over the full duration of the project was met in just the first year of the study. All pilot participants in South Africa consented to have genetic findings related to their child’s NDD returned to the family. Similarly, we observed a high rate (98%) of consent for the option to share cell lines in addition to DNA. Both of these options were available only in South Africa.

We used the University of California, San Diego Brief Assessment of Capacity to Consent (UBACC) to both ensure and measure parents’ understanding of the study prior to consent. As a screening tool, the UBACC is used to identify parents who may require a more comprehensive evaluation of decisional capacity and enhanced consent procedures.^10,11^ Participants need to achieve a score of 14.5 out of 20 to meet the test requirement, with the tool readministered up to a maximum of three trials if necessary, each after additional explanatory efforts. In the first year of data collection, only two parents failed their UBACC administrations, and their families were not included in the study. One of the two parents was reported to have a documented ID. An overwhelming majority of parents showed good understanding of the protocol following detailed explanation by study staff, since only 3% of participants scored below 14.5 on their first trial.^12^

The pilot period was used to further review our data collection strategy and tools. In the Methods section, we share detailed observations about the assessment tools as applied in our context, as well as any adaptations to the tools made at either site. These adaptations were made in response to questions that were contextually inappropriate for caregivers due to cross-cultural differences or linguistic challenges. We also discuss modifications to the protocol, lessons learnt in the implementation of the study, and recruitment strategies used in response to variability in enrollment of participants. For example, in Kilifi, we adapted our recruitment strategy during inclement weather, and in response to challenges recruiting fathers during the work-week. We hope these details will be of benefit to future research projects, and that this suite of pilot results as a whole will encourage more large-scale NDD projects in Africa.

We made one significant addition to the phenotype battery, the Child Behavior Checklist (CBCL)^13^, aiming to strengthen participant behavioral characterization. We initially employed the CBCL to explore the validity of the SNAP-IV in a subset of NeuroDev participants and found it to be of both research and clinical utility. The CBCL, now included in NeuroDev’s core phenotype battery, characterizes a comprehensive assortment of problem behaviors separately in preschool (age 1.5-5 years) and school-age (6-18 years) children. In addition to the research opportunities afforded by the CBCL data, we observed clinical value in use of the CBCL to construct a behavioral profile for children that may need a referral for specialized intervention.

### Phenotypic characteristics of cases ascertained in NeuroDev’s first year

NeuroDev features an uncommonly detailed phenotype battery for a genetic study of its size. In this section, we present phenotypic findings from all 219 cases collected between August 2018 to July 2019. Of those cases, 156 were South African and 63 were Kenyan, reflecting the fact that the Kenyan data collection began halfway through the pilot’s first year; 99 of these 216 cases were included among the exome-sequenced trios (Figure 1A). Overall, cases were 70.2% male, and they ranged in age from 2-18 years (Figure 1B). The majority (80.4%, N = 176) of cases met criteria for global developmental delay or intellectual disability (GDD/ID). Of those with GDD/ID, 91 cases (52% of all GDD/ID cases) also met criteria for autism. An additional 31 cases (14.2% of all cases) met criteria for autism without GDD/ID. A small number (5.5%) of cases did not meet criteria for either GDD/ID or autism, but for other NDDs that are ascertained through NeuroDev: specific learning disabilities, communication disorders, and/or ADHD. The diagnostic composition of cases included in the 99 exome-sequenced trios is highly similar to the full set 219 cases (Figure 1E).

**Figure 1.**
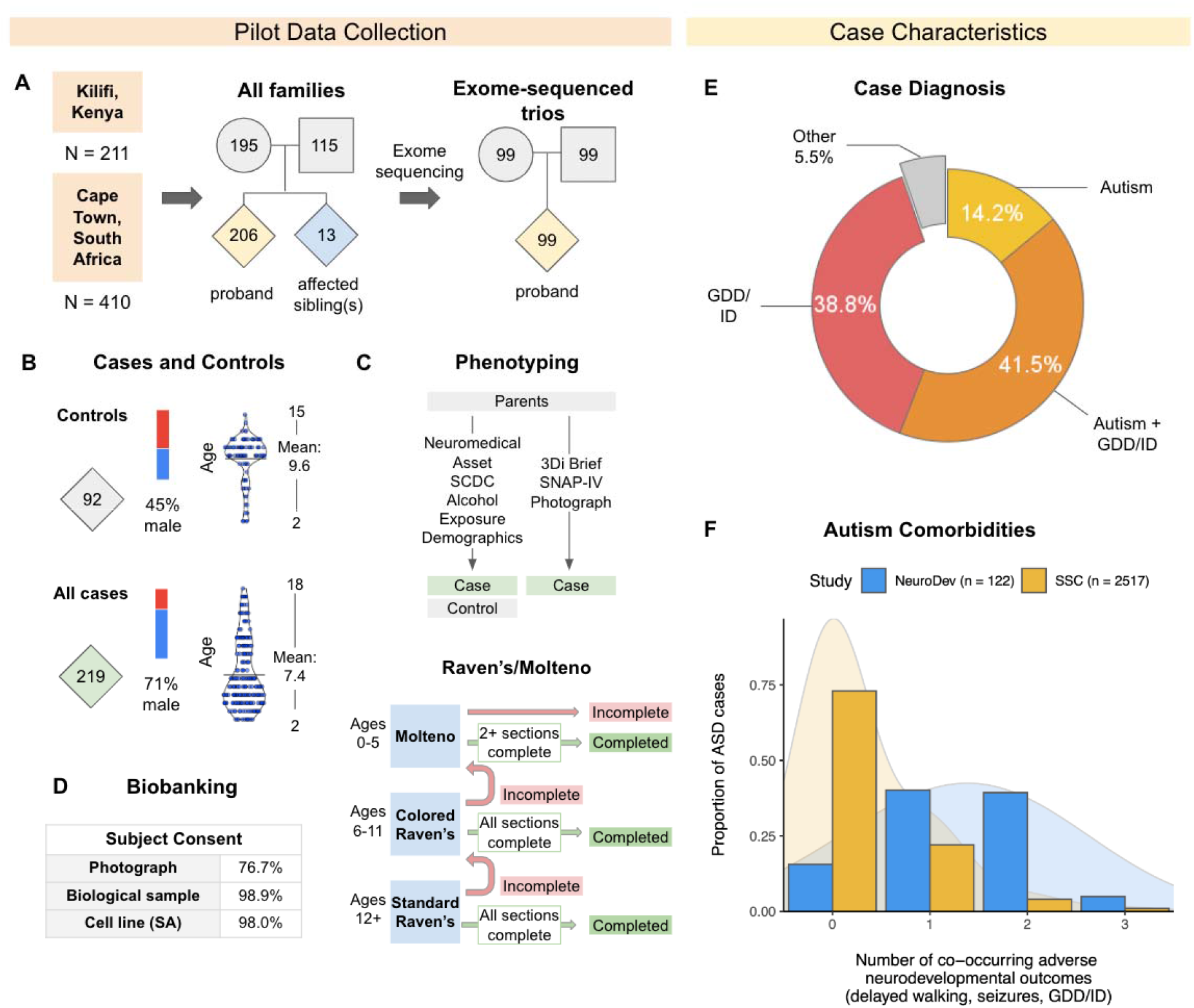
Overview of the NeuroDev Trio Pilot. (A) Data collection and exome sequencing description. (B) Distribution of sex, age, and consent subtypes across cases and controls. (C) Assessments administered to cases and for controls. The flowchart describes Raven’s/Molteno administration processes for those who found completing either test to be challenging. (D) Subject consent rates for biobanking. (E) Diagnostic profiles of the 219 cases in the pilot phase. “Other” diagnoses include specific learning disabilities, communication disorders, and/or ADHD. (F) Number of co-occurring adverse neurodevelopmental outcomes among the NeuroDev and SSC autism cases. Values of 0, 1, 2, or 3 indicate the total number of the following outcomes present: delayed walking, seizures, or GDD/ID. A score of 3 indicates that all three symptoms are present.

The cases were highly ancestrally and linguistically diverse, with more than 40 languages spoken in families and more than 24 ethnicities represented. In keeping with the approach initiated by the 1000 Genomes study, we used language as an indicator of ethnic affiliation on top of self-reported ethnicity. Among cases from Kenya, the majority were from the Mijikenda ethnic group (88.7%) with many of the case families primarily speaking the Mijikenda languages (83.9%) or Kiswahili (14.5%). Within South Africa, most of the cases were of mixed ancestry (45.9%) or identified with multiple ancestries (16.9%), and many others identified as AmaXhosa (13.5%). Reflecting this, the main languages spoken by the case families in South Africa included English (74.3%) and isiXhosa (14.2%). The specific questions on language and ethnicity in the demographic questionnaire were phrased as follows: ‘What is the primary language spoken in [participant’s] home?’ and ‘What is [participant’s] ethnicity or tribe?’

As anticipated, most children with autism in this initial group also met criteria for GDD/ID (74.6%). This is consistent with other descriptions of clinic-based cohorts of children with autism from resource-limited environments.^14–16^ The scarcity of neuropsychiatric specialists and medical resources in the African region contributes to later diagnosis or lack of access to services for children with milder autism symptoms and minimal cognitive impairment.^17–19^

We compared the prevalence of GDD/ID, delayed walking (after 18 months of age) and parent-reported seizures between child cases with autism in NeuroDev and those in the Simons Simplex Collection (SSC), a large United States-based cohort (Figure 1F). In contrast to cases ascertained through the SSC, most NeuroDev autism cases presented with at least one of these co-occurring adverse neurological or neurodevelopmental outcomes: i) seizures, ii) GDD/ID, and/or iii) walking later than 18 months (84.4%). The average number of co-occurring adverse outcomes in the NeuroDev study was 1.34 (N = 122), whereas this number was .33 for SSC (N = 2517, p = 2.2×10^−16^). The difference in the number of co-occurring outcomes likely reflects differences in research setting and case ascertainment, as described above. The rate of co-occurring neurodevelopmental conditions in individuals with autism is associated with average genetic architecture.^20,21^ Most significantly, the case rate of *de novo* protein truncating variants (PTVs) in constrained genes increases with the number of co-occurring adverse neurodevelopmental outcomes. Based on the data in Weiner et al., we conservatively expected a 50% increase in the observed rates of *de novo* PTVs in constrained genes in NeuroDev autism cases relative to SSC and similar cohorts.^21^

The initial 219 phenotyped cases experienced high levels of speech delay, with only 36% of cases meeting criteria for fluent speech. The high rates of delay were expected due to our ascertainment strategy, which included recruitment of cases from neurodevelopmental clinics and special needs schools. Those with a comorbid diagnosis of autism and ID/GDD had the lowest speech level, with 76% of families reporting the use of single words or less. Consistent with this level of developmental delay, many case children also experienced challenges completing the RPM. Among all cases, only 66% of all Standard RPM testers (age 12 or higher) and 44% of all Colored RPM testers (ages 6-11) were able to fully complete age-appropriate versions of the assessment (Supplementary Figure 1, Supplementary Table 1). Among Standard RPM testers that could not complete the assessment, approximately half were able to complete the Colored version of the RPM, and the rest completed the Molteno. Taken together, these findings suggest high severity level of developmental delay in recruited NeuroDev cases, as compared to other NDD cohorts that are widely available.

### Genetic Diversity

To examine genetic diversity, we compared the 99 genotyped trios to individuals from the Human Genome Diversity Panel (HGDP), 1000 Genomes Project (TGP), and the African Genome Variation Project (AGVP). In the context of these global and subcontinental reference panels, we saw that the South African and Kenyan NeuroDev cohorts were ancestrally dissimilar (**Figure 2**). The NeuroDev cohort from Cape Town, South Africa maintained a high proportion of globally admixed individuals, consistent with expectation from previous work ^22^ and maintained a subset of populations lacking out-of-Africa admixture that cluster closely with continental African populations. The NeuroDev Pilot cohort from Kilifi, Kenya was of almost exclusively African genetic ancestry with little evidence of global admixture (**Figure 2A, 2B**).

**Figure 2.**
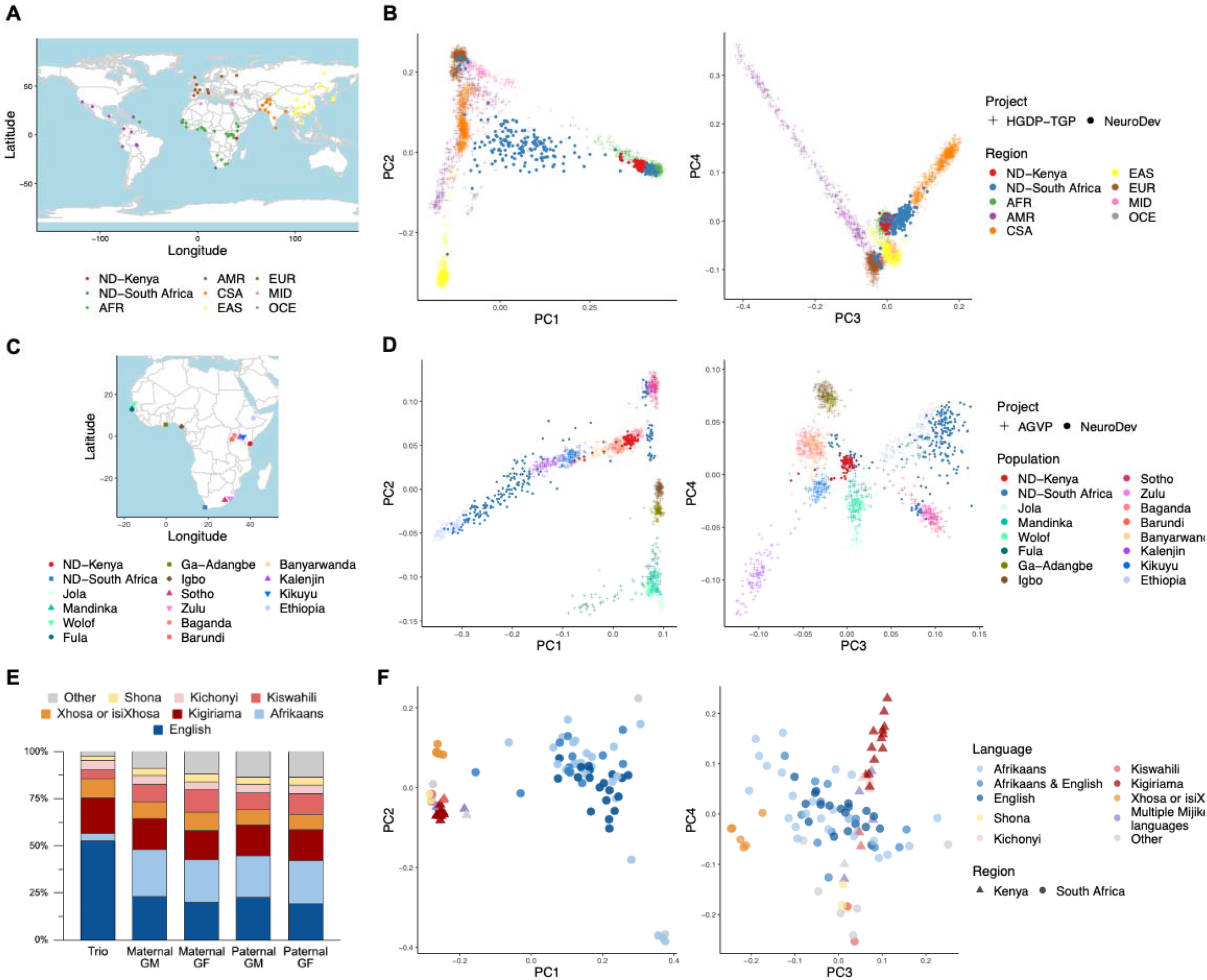
Global and within-Africa population structure of the NeuroDev Trio Pilot. (A) Map of geographic populations represented in the Human Genome Diversity Panel and 1000 Genomes Project (HGDP-TGP) global reference panel. (B) Global PCA plots of the NeuroDev cohort. All NeuroDev trios are projected onto the first 4 PCs of the HGDP-TGP reference panel. (C) Map of geographic populations represented in AGVP, a within-Africa reference panel. (D) African subcontinental PCA plots of the NeuroDev cohort. A subset of sequenced NeuroDev individuals with African genetic ancestry are projected onto the first 4 PCs of the AGVP panel. (E) Ethnolinguistic breakdown of all languages spoken by the 99 genetically sequenced NeuroDev families (“Trio”) and their grandparents (“GF”: grandfather, “GM”: grandmother). “Other” spans a total of 24 languages. (F) PCA of the 99 NeuroDev cases without a reference panel, colored by the respective maternal grandmother’s reported ethnolinguistic background.

Next, we assessed continental African genetic ancestry of NeuroDev cohorts by intersecting the NeuroDev genotyping data with those of populations described in the African Genome Variation Project (**Figure 2C, 2D**).^23^ The first principal component separates Ethiopian populations from other continental African populations due to a back-to-Africa migration and admixture in Ethiopians.^24^ The clustering of some NeuroDev South African samples with these individuals likely indicates out-of-Africa admixture. Individuals of solely African descent in South Africa and Kenya tend to cluster with geographically neighboring subcontinental populations: non-admixed NeuroDev South African samples cluster with the Zulu and Sotho populations of southern Africa; NeuroDev Kenya samples cluster closely with eastern African populations, including Baganda and Barundi ethnolinguistic groups. Given the change in the representation of ethnolinguistic groups over generations (**Figure 2E**), we show that grandparents’ ethnolinguistic groups are interrelated with genetic ancestry, with the maternal grandmother’s spoken languages predicting genetic ancestry of the proband (**Figure 2F**).

### Genetic analyses of the trio pilot data

Of the 99 parent-child trios included in the trio sequencing analysis, 75 were from South Africa and 24 were from Kenya. As of the present analysis, 22 pathogenic or likely pathogenic variants have been identified in those families. We have also identified 7 suspicious variants of uncertain significance (VUS) in emerging disease genes with supportive case data identified through submission to the MatchMaker Exchange (MME). A detailed description of the sequencing and data analysis approach can be found in the STAR Methods. In brief, exome sequencing was performed on each of the trios, and the data was uploaded to the *seqr* platform for analysis.^25^

A total of 13/75 (17.3%) South African cases were solved with pathogenic or likely pathogenic variants in genes with an existing disease association in OMIM.^26^ Of these, 5 cases had a single nucleotide variant (SNV) and 1 case had an insertion or a deletion (indel) (Table 1, Supplementary Table 3). An additional 7 cases had structural variants that included a known OMIM gene (Table 2, Supplementary Table 4). Nearly all of the events were *de novo*, with the exception of one structural variant that was paternally inherited (unknown paternal history of psychiatric diagnoses or NDDs). A total of 9/24 (37.5%) Kenyan cases were solved. Of these, we found 6 cases had a pathogenic or likely pathogenic SNV in genes previously associated with an NDD (Table 1) and 3 cases had a structural variant including a known OMIM gene (Table 2, Supplementary Table 4).

**Table 1.** South African and Kenyan SNV/indel findings.

| Individual | Sex | Clinical Diagnosis | Gene | Variant | Event | Additional Phenotypes of Interest | OMIM # | Variant Interpretation |
| --- | --- | --- | --- | --- | --- | --- | --- | --- |
| South Africa |  |  |  |  |  |  |  |  |
| 859-44206536 | Male | GDD | AGO1 (NM_012199.5) | c.971C>T,<br>(p.Pro324Leu) | missense (MPC<br>= 2.8) | – | – | VUS |
| 859-25391305 | Male | GDD | CREBBP<br>(NM_004380.3) | c.3914+3G>T | extended splice<br>site<br>(CADD = 15) | Craniofacial<br>dysmorphia,<br>abnormalities<br>of the genital<br>system, short<br>stature | #180849 | LP |
| 859-49145346 | Female | GDD | DDX3X<br>(NM_001356.5) | c.599A>G,<br>(p.Tyr200Cys) | missense<br>(MPC = 3.1) | Small for<br>gestational<br>age, abnormal<br>facial shape | #300958 | LP |
| 859-90780619 | Male | GDD | IRF2BPL<br>(NM_024496.4) | c.2137del,<br>(p.Leu713SerfsTer54) | frameshift<br>(CADD = 33) | – | #618088 | LP |
| 859-81577625 | Male | GDD | PPP2R5C<br>(NM_001161725.1) | c.254_259del,<br>(p.Asp85_Phe86del) | inframe<br>deletion<br>(CADD = 17) | Seizure | – | VUS |
| 859-88385997 | Male | GDD,<br>ID | CREBBP<br>(NM_004380.3) | c.5558A>C,<br>(p.Gln1853Pro) | missense<br>(MPC = 1.6) | Hearing<br>impairment,<br>visual<br>impairment,<br>craniofacial<br>dysmorphia,<br>hypertonia | #618332 | LP |
| 859-98643450 | Male | GDD,<br>ADHD | SYNGAP1<br>(NM_006772.3) | c.3795-1G>A | essential splice<br>site<br>(CADD = 35) | Large birth<br>length | #612621 | P |
| 859-21383847 | Male | GDD,<br>autism | SCN2A<br>(NM_001040142.2) | c.2877C>A,<br>(p.Cys959Ter) | nonsense<br>(CADD = 37) | – | #613721 | P |
| 859-33526476 | Female | autism | CACNA1C<br>(NM_001129827.2) | c.4129dup,<br>(p.Arg1377ProfsTer61) | frameshift<br>(CADD = 33) | – | – | VUS |
| 859-76545750 | Male | autism | CACNA1E<br>(NM_001205293.3) | c.3422+1G>A | essential splice<br>site<br>(CADD = 35) | Tall stature,<br>proximal<br>amyotrophy | – | VUS |
| 859-40050374 | Female | autism | MYH10<br>(NM_001256012.3) | c.2555G>A,<br>(p.Arg852Gln) | missense<br>(CADD = 30) | Macrocephaly,<br>high BMI | – | VUS |
| **Kenya** |  |  |  |  |  |  |  |  |
| 860-61235417 | Female | ID | BCL11B<br>(NM_138576.4) | c.1535_1536del,<br>(p.Ala512GlyfsTer4) | frameshift<br>(CADD = 32) | Visual<br>impairment,<br>low BMI | #618092 | LP |
| 860-26955427 | Female | ID | DDX3X<br>(NM_001356.5) | c.1582C>T,<br>(p.Arg528Cys) | missense<br>(MPC = 3.2) | Small for<br>gestational<br>age, muscle<br>weakness | #300958 | LP |
| 860-46028963 | Male | ID | MAPK1<br>(NM_002745.5) | c.952G>A,<br>(p.Asp318Asn) | missense<br>(MPC = 1.4) | Short stature | #619087 | VUS |
| 860-71034936 | Male | ID | TLK2<br>(NM_001284333.2) | c.1655T>C,<br>(p.Leu552Pro) | missense<br>(MPC = 3.4) | Short stature | #618050 | LP |
| 860-73911739 | Female | ID, autism | *DDX3X*<br>(NM_001356.5) | c.894C>A,<br>(p.Cys298Ter) | nonsense<br>(CADD = 24) | Short stature,<br>abnormality of<br>facial<br>musculature | #300958 | P |
| 860-31257300 | Female | ID, autism | *ZBTB18*<br>(NM_205768.3) | c.204_205del,<br>(p.Asp70HisfsTer19) | frameshift<br>(CADD = 29) | – | #612337 | LP |
| 860-33192107 | Male | ID, ADHD | *MBD5*<br>(NM_018328.4) | c.4170G>A,<br>(p.Trp1390Ter) | nonsense<br>(CADD = 41) | – | #156200 | P |
| 860-89059068 | Female | ID, CD | *SF1*<br>(NM_001178030.1) | c.737C>T,<br>(p.Pro246Leu) | missense<br>(MPC = 3.0) | Visual<br>impairment | – | VUS |

**Table 2.** South African and Kenyan structural variant findings.

| Individual | Sex | Clinical Diagnosis | Genomic Region (GRCh38) | Event | Associated Condition | Additional Phenotypes | Variant Interpretation |
| --- | --- | --- | --- | --- | --- | --- | --- |
| South Africa |  |  |  |  |  |  |  |
| 859-10314801 | Male | GDD | chr3:13371737-20095506 | deletion (6.7 Mb) | 3p deletion syndrome (#613792) | Small for gestational age, microcephaly | P |
| 859-97835206 | Female | GDD | chr6:115941808-133892653 | deletion (18 Mb) | Interstitial 6q microdeletion syndrome | Ventricular septal defect, seizure, meningitis, camptodactyly | P |
| 859-44770029 | Female | GDD | chr18:61490305-80247612 | deletion (18 Mb) | Chromosome 18q deletion syndrome (#601808) | Visual impairment, low birth length, abnormal facial shape, hypotonia | P |
| 859-41524687 | Female | GDD | chr22:18985739-21081116 | duplication (2.1 Mb) | 22q11.2 duplication syndrome (#608363) | High birth length, macrocephaly | P |
| 859-50205427 | Female | GDD, autism | chr15:22810652-29822566 | triplication (7.0 Mb) | 15q11-q13 duplication syndrome (#608636) | Small for gestational age | P |
| 859-22089821 | Male | GDD, autism | chr15:30626003-32111997 | deletion (1.5 Mb) | 15q13.3 microdeletion syndrome (#612001) | Small for gestational age, microcephaly | P |
| 859-85638884 | Male | GDD, autism | chr16:29663598-30188229 | deletion (0.53 Mb) | 16p11.2 deletion syndrome (#611913) | – | P |

| Kenya |  |  |  |  |  |  |  |
| --- | --- | --- | --- | --- | --- | --- | --- |
| 860-31775019 | Male | ID | 22:18985739-21081116 | deletion (2.1 Mb) | DiGeorge syndrome (#188400) | Abnormality of facial musculature, seizure | P |
| 860-45665569 | Male | ID | 22:18985739-21081116 | deletion (2.1 Mb) | DiGeorge syndrome (#188400) | – | P |
| 860-94211640 | Male | ID | 22:49883237-50740457 | duplication (0.85 Mb) | 22q13 duplication syndrome (#615538) | – | P |

GDD, global developmental delay; ID, intellectual disability; P, pathogenic. Age at recruitment is reported. All variants were *de novo* except in subject 859-22089821 where the variant was paternally inherited. See Supplementary table 4 for more details.

In addition to the solved cases, 7 variants of uncertain significance (VUS) were considered to be interesting candidate variants based on matches made through the MatchMaker Exchange (MME; Table 1, Supplementary Table 5). By matching cases of similar phenotypic and genotypic profiles, MME provides a rapid, systematic approach to rare disease gene discovery.^27^ The 7 genes containing these variants were not yet listed in OMIM at the time of analysis, and represent emerging gene-disease associations where additional evidence and cases are still needed. All 7 variants will be included in collaborative case series reports on the genes of interest. To date, case studies on three of these genes – *AGO1, CACNA1C*, and *CACNA1E* – have been published.^28–30^ These variants may potentially be reclassified after re-evaluation over time.

NeuroDev participants comprised the only geographic African cases in any of the case series reports, and NeuroDev will be the first African NDD cohort to contribute, at scale, to rare disease discovery activities. As described by the NHGRI Atlas of Human Malformations initiative, syndromic NDDs often vary in their phenotypic presentation between ancestral groups^31^, a phenomenon particularly well documented with regard to canonical facial features. Further analysis of how phenotypic features differ between ancestral groups will help clarify which features are in fact “core” to a genetic syndrome, and which vary based on ancestral group or environment.

## Discussion

People of African ancestry have been grossly underrepresented in genetic studies, across domains and disciplines.^34,35^ In aggregate, this is most visible through the constitution of large genetic databases like gnomAD, in which only 14% of individuals (N ≈ 27,000) have some African ancestry.^36,37^ As no sequenced cohorts from the African continent are currently present in gnomAD, the great majority of those 27,000 individuals are American, and typically have a mixture of West African and European ancestry.^38,39^ If individuals of African ancestry remain underrepresented in genetic research, they will continue to be less likely to receive accurate genetic diagnoses and less likely to benefit from advances in genomic science and medicine.^40–43^

The NeuroDev project was built to address this representation gap and its scientific, medical, and ethical consequences. Those living in Africa have greater genetic variability than any other human ancestral group, rendering their underrepresentation a substantial barrier to human genome characterization and scientific equity.^34^ There are many reasons that NDD collections have historically been conducted in the United States and Western Europe, chief among them better access to project funding and infrastructural support. The lack of precedent produced common concern about NeuroDev’s feasibility, particularly with regard to the planned case collection rate, and the ambitious case characterization schedule. We hope that the results described here will increase the research community’s confidence in large scale data collection efforts in underrepresented populations, and that many other studies will be able to contribute to greater African representation in NDD genomics.

We also hope that the high rate of participant family interest in receiving genetic results will shed light on the need for consistent access to genetic testing services. To date, all NeuroDev participants in South Africa readily consented to have genetic findings related to their child’s NDD reported back to them by a clinician. Similarly, NeuroDev South Africa observed a very high (98%) rate of consent to generation and sharing of stem cell lines in addition to sharing DNA. This provides us with the opportunity to contribute to much needed diversity to global stem cell collections, which are also heavily biased towards European ancestry.^44^ This high degree of participant enthusiasm in South Africa has led to investigations of similar possibilities in Kenya. The NeuroDev Kenya team has recently been funded by a Fogarty award to support, in part, community engagement on the ethics of cell line generation for the cohort in Kilifi.

While NeuroDev provides in-depth phenotypic characterization of its subjects, this data may still reflect regional differences. Kilifi is a rural coastal town with an agriculture-based economy, with an absolute poverty rate of 46.4% and limited access to school-based education.^45^ In contrast, Cape Town is a large urban setting and is among the wealthiest cities in Africa, conferring relative educational and resource advantages. As many behavioral measures are known to be tied to socioeconomic status or maternal education, our behavioral outcomes likely reflect differences in regional context. Thus, we encourage caution when making comparisons of the outcomes of phenotypic assessments, such as RPM scores, across the two sites.

The four-year target sample for NeuroDev is 1800 cases, 2100 parents of cases and 1800 controls, the largest study to date investigating NDDs on the African continent. The NeuroDev project will, in full, create a public resource for medical genetics research that includes, for thousands of African individuals: genome-wide common variant data; exome sequencing data; comprehensive phenotypic data including detailed cognitive, behavioral, and health information; cell lines (lymphoblastoid and induced pluripotent cell lines); and photographs capturing dysmorphic features. This critically needed line of work will help address the scientific, medical, and ethical consequences of the genomic research representation gap.

## STAR Methods

### NeuroDev Protocol and Informed Consent Procedures

NeuroDev’s information and consent document is the culmination of a series of focus groups and meetings across both sites that were conducted to help develop the language of the document so that it could communicate complex scientific terms in a manner accessible to research participants who may not have completed formal schooling. There were three focus groups in total. The first included parents of children with NDDs, the second included community leaders from NGOs, centers, schools, etc. The third included healthcare workers who work in clinics with NDDs. The key outcomes from the focus group meetings were: Firstly, that lengthy detailed explanations hindered understanding of the scientific terms. Participants tended to be more confused and overwhelmed by lengthy explanations. Therefore, information in the consent document is divulged in ways that keep descriptions short and concise. Secondly, the use of visual examples to help explain DNA were very helpful. For example, pointing out physical similarities between family members as a way of illustrating the inheritance of DNA. Thirdly, it is easier for participants to understand concepts if examples are used that participants are familiar with. For example, when explaining that the information gathered in our research can be used for other purposes. We provide examples of well-known diseases such as diabetes and cancer. This helped participants easily relate to the conveyed concepts. In Kilifi, there was an emphasis on community engagement activities involving discussing what genes are and how variation in genes is associated with neurodevelopmental disorders. This was carried out during meetings with parents of children with neurodevelopmental disorders, teachers and community members.

After an information sharing session and before potential participants consent to enrolment into the study, the UBACC is administered. The UBACC creates an opportunity for discussion with participants around aspects of the protocol they may not have grasped when the study is first explained to them. It has also shown to be effective in identifying participants who may not fully comprehend the protocol. The South Africa Team have had a small number of failures on the test. These were based on 1) the language of the test (i.e. French) and 2) comprehension difficulties of the participant (one participant had ID, and the other had a history of strokes and depression).

The novelty of genetic research in the African context means that the study is important for its potential benefit to African medical and research communities. It is, by necessity, also precedent-setting. There is a lack of precedent around important issues, such as the return of incidental findings to our participants and lack of qualified personnel such as genetic counselors in many parts of sub-Saharan Africa. The evidence base to support feedback of most incidental genomic findings for African populations is not robust. There is virtually no empirical data describing relevant African stakeholders’ preferences and perspectives, including research participants, ethics committee members, researchers, and research regulators on these issues. The ethics arm of the NeuroDev study is currently investigating this and will add evidence on this pertinent issue.

### NeuroDev Data Pipeline

Data is collected on tablets using the REDCap Mobile App during assessments. Every participant is assigned a subject ID and has an intake form filled out. Based on this information, branching logic programmed into REDCap determines which tools and fields populate for the assessor to complete. At the end of an assessment day, data from the tablets is uploaded to the NeuroDev project on the REDCap servers. The team at the Broad Institute then uses the *scred* package – a Python implementation of the REDCap API – to pull raw records by subject ID and assign codes to missing data to differentiate between truly missing fields and non-applicable fields based on the REDCap branching logic. A Python sync engine then organizes the record data by assessment tool, scores assessments like the Raven’s Progressive Matrices and 3Di, and pushes it to a MySQL database hosted on the Google Cloud. The sync procedure is generally run on a weekly basis to keep the database current. All summary and quality-control (QC) reporting is derived from either direct SQL queries to the database or export to pandas dataframes for analysis with Python scripts.

The QC reporting process relies on querying data from the Broad Database and REDCap logging files to produce a comprehensive report of missing, overwritten, or inconsistent participant data. Run on either a date range or a list of specific subject IDs, the program produces summary reports on any of five distinct metrics: completion of REDCap forms, missing required data fields (regardless of whether a form was marked complete), comments entered by assessors at the sites, logical cross-checks on completed data, and fields found to be overwritten in the REDCap log. Subject and site IDs as well as interview times and assessor names are included on each of these reports so the Broad team can easily identify both single issues and larger patterns. Run and reviewed weekly, the program improves data visibility and allows consistent feedback to the sites on improving or correcting collection procedures.

### NeuroDev Subject Recruitment

NeuroDev participants were recruited from two sites, Red Cross War Memorial Hospital in Cape Town, South Africa and KEMRI-Wellcome Trust Research Program in Kilifi, Kenya. Participants were recruited from previous studies, specialized clinics and special schools in Kilifi County, Kenya. For the resampling of participants of earlier studies, the team focused on children about to cross the 18-year-old threshold into adulthood. Participants in Cape Town were recruited from developmental clinics using a combination of clinician referral, waiting room flyers and adverts, and cold recruitment from the Red Cross Memorial Children’s Hospital Department of Pediatrics and Child Health Neurodevelopmental Clinic. Cases and affected siblings included in the study had clinical diagnosis of a neurodevelopmental disorder, were within the specified age range (2-18 years old), and willing to participate. Cases were excluded if they had a co-occuring primary neuro-motor condition such as cerebral palsy or Downs Syndrome. Controls were included in the study if they did not have a diagnosis of a neurodevelopmental disorder, were within the study age range and were matched according to catchment area, ancestry and age. In Kenya, the recruitment of controls also drew from the Kilifi health demographic surveillance system (KHDSS) cohort. The KHDSS, which has been active since 2002, surveys a representative community sample every three months. Control participants in Cape Town were recruited from various outpatient clinics at the Red Cross War Memorial Children’s Hospital.

Written consent was sought for participation of the child controls and child cases from their parents or caregivers, and additional consent was sought from parents of the cases for their own participation in the study. Ethical approval was sought in the site institutional review boards as well as at the Harvard T.H Chan School of Public Health. In Kilifi, Kenya, approval was granted by the Scientific Ethics and Review Unit (KEMRI/SERU/CGMR-C/104/3629) and Health Research Ethics Committee (HREC REF:810/2016) in Cape Town, South Africa.

### NeuroDev Phenotypic Data Collection

#### 1. Assessments

##### Molteno Adapted Scales

The Molteno has proven to be quite effective in helping us capture the neurodevelopmental profile of children under six years of age. The test’s flexible approach to item administration and incidental observations has allowed us to test and score the behavior of even the most non-compliant children. The Molteno had not been validated and adapted for use in Kilifi. We pre-piloted the Molteno, recruiting a random subset of 100 children between 2-5 years in the community and their parents. A small number of items have had to be adapted to make them more appropriate for our population, e.g., “child has a tricycle at home” was changed into “child can walk on his/her tiptoes” along with the administration of a few fine-motor related items.

##### Swanson, Nolan and Pelham (SNAP) ADHD-IV Rating Scale

The SNAP was insightful and was straightforward in its administration in child cases over six years of age. But it presented some limitations for preschool-aged children. Specifically, some items refer to homework and other tasks that would be cognitively challenging and therefore not a representative skill for children who are not yet of school-going age. The instructions for the test administration did not allude to a lower age limit. After a review of the test literature, it was determined that the test was not suitable for children under six years of age and would only be administered to children over the age of 6 years.

##### *Child Behavior Checklist* (CBCL)

The CBCL was implemented into the NeuroDev protocol to help validate the SNAP. This would form a central component of a Master’s research project. Because of the limited utility within the broader aims of the protocol, the test was initially piloted, and the utility would later be evaluated. The CBCL was kept on beyond the pilot stage because it proved to be valuable within the clinical context in both sites as it captures clinically rich information that will help phenotype our population that other tests in the phenotypic battery might not, e.g. attention in children under six years, which was lost to the study when the SNAP was amended. The data from the test will also be beneficial to two doctoral students attached to NeuroDev.

When the NeuroDev Kenya team began administering the CBCL, it was noted there were some questions that parents struggled with when giving responses and requested clarification of the questions. Some of the questions that have come up as needing elaboration included: Argues a lot, Fails to finish things he/she starts, There is very little he/she enjoys, Bragging, boasting, Can’t get his/her mind off certain thoughts; obsessions, Confused or seems to be in a fog, Daydreams or gets lost in his/her thoughts, Strange behavior, Strange ideas, Wishes to be of the opposite sex.

The NeuroDev Kenya team discussed this and came up with a few changes in the assessment flow. The team started the behavioral assessment battery with the CBCL and thereafter the 3Di, SNAP, SCDC. It was noted that the 3Di, which includes standard examples, could have primed the caregivers to expect some examples. The team also added the prompt that apart from the aim of the assessment being exploring children’s behavior: “This assessment describes behaviors of children from a wide range of ages (6-18 years) as such you might find that some questions may seem unsuitable, we would still like to hear from you, to be best of your knowledge, the extent to which your child may have these behaviors.”

##### Developmental Dimensional and Diagnostic Interview (3Di)

The 3di has proven to be a concise evaluative tool for the presence of autistic features in children. It is well-received and requires only minor additional probing for its questions. We have found that, in both the South African and Kenyan context, parents with children who have been diagnosed with autism often go through the questionnaire with ease, likely because of their familiarity with the item constructs. In contrast, parents of children who do not demonstrate autistic behaviors generally require more probing, possibly because they are less familiar with the item constructs.

##### Raven’s Progressive Matrices

The RPM has generally been well-received by our participants; however, we have noticed that on a few occasions participants will make errors in the initial teaching items which suggest that this method of reasoning, i.e., deductive reasoning, is more novel within our context than the test has accounted for. For instance, some participants may choose to select their responses based on matching the features of the response items to those in the stimulus box, instead of deducing properties of the missing image, based on the pattern in the stimulus box. This type of error turns the test into a perceptual test for some participants (at least initially) instead of a test of deductive reasoning.

##### Blood draw

The blood draw can be a discomforting experience of data collection – for the research team, the parents and, most importantly, the child. The management strategy of each blood draw takes account of the idiosyncrasies of each child, in order to reduce any potential distress, time, and the possibility of injury. The teams have found that having a frank and upfront conversation with the parent(s) of the child about what to expect and to determine how they feel their child will respond, is best. Some children will respond best if they know what will happen. The South African team developed “social stories”; a visual aid tool that explains each step of the process to the child using simple language and pictures. The story is written in the first-person, from the point of view of the child, helping the child visualize the experience. The Kenya team adapted and translated the social stories for use in their context as well. We conducted a saliva pilot study on a subset of participants from whom saliva was obtained in addition to a blood sample. Of the 131 samples, 97.15% of DNA extracted passed sample QC, indicating that saliva would be a strong option for DNA in future cases.

##### Photos

Participants in South Africa appear to be most apprehensive about having their children’s photos taken. As of December 2019 76%, of participants have consented to having their photo taken. Participants often report wanting to protect their children’s privacy and preventing their children’s images falling into the hands of unscrupulous characters as reasons for not consenting to the photos. In Kenya, the use of photos was a point of concern during the approval process, as a result, the use of photos for publication and other public use is a twofold process. During the consenting process, participants are informed that the taking of photos is a core part of the assessment process and that before a photo is shared to other research teams, that we will re consent them to get permission to use the selected photos.

##### Cell lines and stem cells in South Africa

The South Africa team has observed a much higher rate of consent for cell lines than previously anticipated. As of December 2019, 96% of participants had consented to cell lines. The Kenya team has not implemented cell line and stem cell collection. The ethics arm of the study is collating views from community members and key stakeholders on collection of cell lines and will present these findings in the coming years.

#### 2. Case ascertainment

##### Case ascertainment In Kenya

Intellectual Disability is more readily ascertained in Kenyan Special Schools through the Educational Assessment and Resource Centers, however, autism diagnosis is not as readily ascertained, especially when comorbid with another NDD (Abubakar et al., 2022). As some of the Kenyan cases are recruited from Special Schools, to ensure we do not miss cases with autism, we began clinically assessing children that came in with only a reported diagnosis of ID, but who in the course of the NeuroDev assessments battery presented with many autism traits (i.e., they endorse a lot of the symptoms on the 3Di). Using their clinical judgment and a general endorsement pattern on the 3di, in particular the repetitive behaviors section questions, the case was further assessed by an experienced clinician on the team and a psychologist using the DSM-5 checklist and clinical judgment to confirm whether the child has autism.

##### Case ascertainment in South Africa

Children with a known NDD diagnosis were recruited from the neurodevelopmental and genetic outpatient clinics from Red Cross War Memorial Children’s Hospital and Tygerberg Hospital in Cape Town, South Africa. Eligible cases were identified through clinician referral. Additionally, families in the clinic waiting rooms were approached and screened for eligibility and invited to take part in the study. Both hospital clinics are specialist developmental outpatient clinics, which are led by relevant sub-specialists (Paediatrics, Paediatric Neurology, Medical Genetics and Developmental Paediatrics). Children enrolled in the study have been formally diagnosed by these teams through standardized clinical assessments and benchmarked against the DSM-5 criteria.

#### 3. Lessons Learned

##### Reflection on parent-recalled dates in Kenya

The Kenyan cohort is generally older in comparison to the South Africa cohort, as such recall of certain milestones and ages can be challenging for the caregivers. The team uses anchoring when trying to help parents answer an age-related question. First using age bands: Under 1 year, 12-18 months (6 months after a year), 2 years (how many months before or after?). Thereafter, they also employ the use of cultural events and family events to enhance the age anchoring technique.

##### General approach to data collection

The case children are highly variable in terms of their type of neurodevelopmental disorder. As such there is great variation in how each family approaches dealing with their child’s condition. The researchers have adopted a series of principles that encourages sensitivity and reflexivity to the needs and expectations of each family during data collection. These include maintaining a sense of calm and patience during testing to reduce collective anxiety; being respectful of each family’s journey and process; taking the time to explain what is going on to the child, even if they are non-verbal and look as if they don’t understand what you’re saying; trust and rely on each other as a team and being open to learn from each other; and ensure the researchers, the participants, and their families are ready for the blood draws as they can be very traumatic.

##### Follow-up after scheduling of assessments

Throughout the study, the teams incorporated intensive follow-up efforts to increase the rate with which fathers participated in the study, to increase the trio collection. They were followed-up regularly and allowed flexible scheduling, sometimes during other data collection slots (this was possible because only one research nurse/clinician was needed to complete data collection of returning fathers: including consenting, cognitive testing and blood draw) to accommodate logistical reasons for not attending data collection. In Kilifi, fathers are often away at work during the day, when the recruitment team does the home visit, and work most of the week and may not be able to get time off work. To work around this, the study team arranged for ‘Father’s Day’ Data collection days on Saturday.

#### Phenotypic Data Analysis Methods

Self-reports of ancestry and language were collected as part of the Demographics tool on REDCap. To assess developmental or intellectual delays, we administered either the Molteno or the Raven’s Progressive Matrices (RPM) to their respective age groups and measured rates of completion. Successful completion of the Molteno was defined as the completion of at least 2 of the 4 domains implicated in the assessment. Completion of the RPM was defined as being able to complete all questions on the assessment. Proportions of testers who successfully completed the respective tests at age ranges of 0-5 (Molteno), 6-11 (Colored RPM), and greater than 12 (Standard RPM) was computed and reported. For subjects who could not complete age-appropriate assessments, the rates of completion of tests below their age level was reported for each age group.

Neurodevelopmental diagnosis of cases was collected through the Neuromedical Assessment tool. The cases were placed into four distinct categories: autism, GDD/ID, autism and GDD/ID, and other diagnoses (which included ADHD, communication disorder, and specific learning disorders). Children that met DSM-5 criteria for either GDD or ID or were diagnosed with at least borderline delay from the Molteno were included in the GDD/ID category. For each of these four categories, we assessed the proportions of those with fluent speech levels. Speech fluency was determined by the 3Di, which assesses the child’s speech on a scale of no words, single words, multiple words, and fluent speech based on parent interview.

The Simons Simplex Collection (SSC) data set was used to compare phenotypic outcomes of the 122 cases with autism ascertained in NeuroDev. The SSC consists of a deeply phenotyped sample of more than 2500 families with a child diagnosed with autism in the United States.^5^ We grouped cases with autism from both datasets based on the number of adverse co-occurring neurological and developmental outcomes, including ID, a positively associated history of seizures, and motor delays (defined as either a gross motor diagnosis from the Molteno, or an age at first steps higher than 18 months). In SSC, ID was defined as cases with an IQ < 70. From the symptom distributions within each study, the average number of co-occurring symptoms was computed, and the difference between these means was assessed using a Mann-Whitney U test.

#### Exome Sequencing & Data Processing Methods

Data generation and analysis were done in collaboration with the Broad Institute Center for Mendelian Genomics, with sequencing performed at the Genomics Platform and data processing at the Data Sciences Platform of the Broad Institute of MIT and Harvard. Libraries from DNA samples were created with a Twist exome capture (37 Mb target) and sequenced (150 bp paired reads) to >85% of targets at >20x, comparable to ∼55x mean coverage. Sample identity quality assurance checks were performed on each sample. The exome sequencing data was de-multiplexed and each sample’s sequence data aggregated into a single Picard CRAM file. Exome data was processed through a pipeline based on Picard, using base quality score recalibration and local realignment at known indels, aligned to the human genome build 38 using BWA, and jointly analyzed for single nucleotide variants (SNVs) and insertions/deletions (indels) using Genome Analysis Toolkit (GATK) Haplotype Caller package version 4.0.10.1.

After variant calling, sex, ancestry and relatedness to other samples were inferred using the CMG sample QC pipeline and compared to sample metadata to identify and correct sample swaps, basic functional annotation was performed using Variant Effect Predictor (VEP). The joint variant call file was then uploaded to the *seqr* platform and imported to Hail for further annotation and analysis.

Copy-number variants (CNVs) were discovered from the exome sequencing data following GATK-gCNV best practices. Read coverage was calculated for each exome using GATK CollectReadCounts. After coverage collection, all samples were subdivided into batches for gCNV model training and execution; these batches were determined based on a principal components analysis (PCA) of sequencing read counts. After batching, one gCNV model was trained per batch using GATK GermlineCNVCaller on a subset of training samples, and the trained model was then applied to call CNVs for each sample per batch. Finally, all raw CNVs were aggregated and post-processed using quality- and frequency-based filtering to produce the final CNV callset.

#### Exome Sequencing Data Analysis Process

Upon completion of data generation, both the SNV/indel and CNV callsets were uploaded to *seqr*, the centralized genomic analysis platform used by the Broad Institute’s Center for Mendelian Genomics (CMG), for clinical genetics analyses. The CMG analysis team deployed a standard analysis protocol across all NeuroDev trios to identify potential disease-causing variants. The first round of analysis consisted of a review of variants from a *de novo*/dominant and a recessive search. The *de novo*/dominant search filters for variants present in affected family members and absent from unaffected family members. The variant types returned from this standard search include deletions, duplications, protein truncating variants, and missense variants that have an allele frequency <0.1% across population databases (gnomAD v2/v3/SV,

1000 Genomes, ExAC, and TopMed) and <1% in the CMG internal rare disease dataset, that pass QC, and that have a GQ ⋝20 and an allele balance ⋝0.2. The recessive search returns homozygous recessive, compound heterozygous, and X-linked recessive variants in affected individuals, considering phasing for any available parents. It searches for biallelic variants across deletions, duplications, protein truncating variants, or missense variants that have an allele frequency <1% across population databases and <3% in the CMG internal rare disease dataset, that pass QC, and that have a GQ ⋝20 and an allele balance of ⋝0.2. If candidate variants were not identified after the first round of analysis, the search criteria was adjusted to include additional variant annotation types (synonymous variants, extended splice site variants, and 5’ and 3’ UTR variants), and quality parameters were relaxed to allow for the review of indels that did not pass QC.

Potential causal variants were subjected to rigorous evaluation of the evidence for pathogenicity following criteria established by the American College of Medical Genetics and Association for Molecular Pathology.^46^ The study’s primary focus was on well-established disease genes, using information drawn from a variety of sources such as OMIM. The phenotype data of NeuroDev participants was assessed for possible consistencies with the previously reported clinical presentations associated with known disease genes, bearing in mind potential deviations from expectation due to ancestry.

Genes that are not yet associated with a well-established human disease in OMIM were carefully evaluated using a variety of sources of evidence, including constraint scores, transcript and protein expression databases, model organism data, and a review of the available literature. All variants identified in candidate disease genes were entered into the Matchmaker Exchange (MME) network through *seqr* in order to identify additional cases with overlapping phenotypes and variants in the same candidate genes, to help better characterize potential novel gene-disease relationships.

#### Genotyping and Analysis Process

Genotyping was performed on all samples using the Illumina Infinium Global Screening Array (GSA) at the Genomics Platform at the Broad Institute, which is supported by a LIMS-tracked, automated processing system that utilizes various sample handling robots and Illumina iScans to generate intensity data from the arrays. After scanning, the data is processed through our automated genotype calling pipeline. The platform releases raw data files (idats, gtc) as well as VCF, and allows for the conversion to PLINK ready called genotype files (ped/map) generated using two different calling algorithms: Illumina GenCall (Autocall) for common variants, and zCall for rare variants including the custom exome/clinical content.

We applied the GWASpy pipeline (https://github.com/atgu/GWASpy) pre-imputation QC module to perform variant-level QC. We used default filters, including MAF, call rate, Mendelian error rate, sex checks, inbreeding coefficient, and Hardy-Weinberg disequilibrium. To visualize genetic diversity across NeuroDev samples, we also performed a principal component analysis (PCA) on array data. The PCA module in GWASpy applied LD-pruning, relatedness estimates, and other necessary filters. We used the joint Human Genome Diversity Project and 1000 Genomes Project reference as a global reference, and the African Genome Variation Project as the subcontinental reference in our population PCA plots. We performed projection PCA, in which the reference data is used to define the principal components and NeuroDev data is projected onto that space, to prevent relatedness effects from our family-based data.

## Supporting information

Supplementary Tables and Figure File_NeuroDev Trio Pilot

## Data Availability

The trio data presented here can be accessed through National Human Genome Research Institute (NHGRI) Analysis Visualization and Informatics Lab-space (ANVIL) controlled access data repository.

https://anvilproject.org/data/

## Supplementary Tables and Figures

Supplementary Table 1: Case data completeness for trio pilot Supplementary Table 2: Parent data completeness for trio pilot

Supplementary Table 3: Reportable SNV/indel pathogenic and likely pathogenic findings in known OMIM disease genes

Supplementary Table 4: Structural variant pathogenic and likely pathogenic findings Supplementary Table 5: Variants of unknown significance in novel candidate genes Supplementary Figure 1: Flowchart of administration and completion rates for the Raven’s Progressive Matrices and Molteno assessments

Supplementary Figure 2: Breakdown of represented ethnolinguistic groups among the 219 NeuroDev cases

## Acknowledgements

We are extremely grateful to the family members for participating in this research. We are grateful to the clinical laboratories and biobank teams at KEMRI-Wellcome Trust and the University of Cape Town, the community liaison group and neuro-epilepsy clinic at the KEMRI-Wellcome Trust and developmental and allied clinics at Red Cross Memorial Hospital. We acknowledge James Swanson, Edith Nolan and William Pelham and team for the use of the SNAP-IV; David Skuse, Richard Warrington, Will Mandy and team for the use of the 3Di and SCDC; the ASEBA team for the use of the CBCL; Christopher Molteno and team for the use of the Molteno Adapted Scales; and the John C Raven, John H Court, and team for the use of the Raven’s Progressive Matrices.

## Author Contributions

E.R, K.A.D, A.A, C.N designed the study. The NeuroDev Project members, including P.K, B.C, E.E, M.K, A.N, B.M, S.M, K.M, were involved in data collection, H.A.K, E.O, J.A, S.Br, N.B, C.K, P.M, B.M were involved in the curation and analysis of the data, P.K, H.A.K, B.C, E.O, E.R wrote the manuscript with input from all authors, P.K, B.C, E.E, A.G, K.M were involved in project administration and C.AT, S.Ba, H.B, A.L, D.G.M, A.S.J, M.B.S, M.E.T, VdM, A.M, L.L.N, CvdM, C.N, A.O.L, A.A, K.A.D and E.R supervised various aspects of the project and the core project teams.

The NeuroDev Project members (current and previous) in addition to the named authors include: Aleya Zulfikar Remtullah, Alex Macharia, Ann Karanu, Carmen Swanepoel, Claire Fourie, Constance Rehema, Deepika Goolab, Dorcas Kamuya, Dorothy Chepkirui, Este Sauerman, Eunice Chepkemoi, Fagri February, Fatima Khan, Felicita Omari, Gina Itzikowitz, Javan Nyale, Jantina de Vries, Jess Ringshaw, Johnstone Makale, Judy Tumaini, Kegan Chase, Lizette Rooi, Moses Mangi, Moses Mosobo, Megan Page, Nicole Mciver, Nonceba Ngubo, Pauline Samia, Phatiswa Lopoleng Ranyana, Racheal Mapenzi, Rachel Odhiambo, Raphaela Itzikowitz, Rene Lepore, Rizqa Sulaiman-Baradien, Samuel Mwasambu, Shaheen Sayed, Susan Wamithi, Tabith Shali, Thandi Xintolo, and Zandre Bruwer.

## References

1. de Menil V, Hoogenhout M, Kipkemoi P, Kamuya D, Eastman E, Galvin A, et al. The NeuroDev Study: Phenotypic and Genetic Characterization of Neurodevelopmental Disorders in Kenya and South Africa. Neuron. 2019 Jan 2;101(1):15–9.

2. Skuse DH, Mandy WPL, Scourfield J. Measuring autistic traits: heritability, reliability and validity of the Social and Communication Disorders Checklist. Br J Psychiatry. 2005 Dec;187(6):568–72.

3. Skuse D, Warrington R, Bishop D, Chowdhury U, Lau J, Mandy W, et al. The Developmental, Dimensional and Diagnostic Interview (3di): A Novel Computerized Assessment for Autism Spectrum Disorders. J Am Acad Child Adolesc Psychiatry. 2004;43(5):548–58.

4. Swanson JM, Schuck S, Porter MM, Carlson C, Hartman CA, Sergeant JA, et al. Categorical and Dimensional Definitions and Evaluations of Symptoms of ADHD: History of the SNAP and the SWAN Rating Scales. Int J Educ Psychol Assess. 2012 Apr;10(1):51–70.

5. Fischbach GD, Lord C. The simons simplex collection: A resource for identification of autism genetic risk factors. Neuron. 2010;68(2):192–5.

6. Robinson EB, Samocha KE, Kosmicki JA, McGrath L, Neale BM, Perlis RH, et al. Autism spectrum disorder severity reflects the average contribution of de novo and familial influences. Proc Natl Acad Sci. 2014 Oct 21;111(42):15161–5.

7. Raven J. The Raven’s progressive matrices: change and stability over culture and time. Cogn Psychol. 2000 Aug;41(1):1–48.

8. Charman T, Jones CRG, Pickles A, Simonoff E, Baird G, Happé F. Defining the cognitive phenotype of autism. Brain Res. 2011 Mar 22;1380:10–21.

9. DiStefano C, Sadhwani A, Wheeler AC. Comprehensive Assessment of Individuals With Significant Levels of Intellectual Disability: Challenges, Strategies, and Future Directions. Am J Intellect Dev Disabil. 2020 Nov 19;125(6):434–48.

10. Jeste DV, Palmer BW, Appelbaum PS, Golshan S, Glorioso D, Dunn LB, et al. A new brief instrument for assessing decisional capacity for clinical research. Arch Gen Psychiatry. 2007 Aug;64(8):966–74.

11. Seaman JB, Terhorst L, Gentry A, Hunsaker A, Parker LS, Lingler JH. Psychometric Properties of a Decisional Capacity Screening Tool for Individuals Contemplating Participation in Alzheimer’s disease Research. J Alzheimers Dis JAD. 2015;46(1):1–9.

12. Campbell MM, Susser E, Mall S, Mqulwana SG, Mndini MM, Ntola OA, et al. Using iterative learning to improve understanding during the informed consent process in a South African psychiatric genomics study. PLOS ONE. 2017 Nov 29;12(11):e0188466.

13. Achenbach TM, Dumenci L, Rescorla LA. Ratings of relations between DSM-IV diagnostic categories and items of the CBCL/6-18, TRF, and YSR. Burlingt VT Univ Vt. 2001;1–9.

14. Bakare MO, Ebigbo PO, Ubochi VN. Prevalence of autism spectrum disorder among Nigerian children with intellectual disability: a stopgap assessment. J Health Care Poor Underserved. 2012 May;23(2):513–8.

15. Mpaka DM, Okitundu DLEA, Ndjukendi AO, N’situ AM, Kinsala SY, Mukau JE, et al. Prevalence and comorbidities of autism among children referred to the outpatient clinics for neurodevelopmental disorders. Pan Afr Med J [Internet]. 2016 Oct 17 [cited 2020 Nov 25];25. Available from: https://www.ncbi.nlm.nih.gov/pmc/articles/PMC5324163/

16. Bello-Mojeed MA, Omigbodun OO, Bakare MO, Adewuya AO. Pattern of impairments and late diagnosis of autism spectrum disorder among a sub-Saharan African clinical population of children in Nigeria. Glob Ment Health. 2017 Mar 21;4:e5.

17. Hahler EM, Elsabbagh M. Autism: A global perspective. Curr Dev Disord Rep. 2015;2(1):58–64.

18. Mazurek MO, Handen BL, Wodka EL, Nowinski L, Butter E, Engelhardt CR. Age at First Autism Spectrum Disorder Diagnosis: The Role of Birth Cohort, Demographic Factors, and Clinical Features. J Dev Behav Pediatr. 2014 Dec;35(9):561–9.

19. Ruparelia K, Abubakar A, Badoe E, Bakare M, Visser K, Chugani DC, et al. Autism spectrum disorders in Africa: current challenges in identification, assessment, and treatment: a report on the International Child Neurology Association Meeting on ASD in Africa, Ghana, April 3-5, 2014. J Child Neurol. 2016;31(8):1018–26.

20. Liao C, Moyses-Oliveira M, Esch CED, Bhavsar R, Nuttle X, Li A, et al. Transcriptional patterns of coexpression across autism risk genes converge on established and novel signatures of neurodevelopment [Internet]. medRxiv; 2022 [cited 2022 Mar 8]. p. 2022.02.28.22271620. Available from: https://www.medrxiv.org/content/10.1101/2022.02.28.22271620v1

21. Weiner DJ, Wigdor EM, Ripke S, Walters RK, Kosmicki JA, Grove J, et al. Polygenic transmission disequilibrium confirms that common and rare variation act additively to create risk for autism spectrum disorders. Nat Genet. 2017 Jul;49(7):978–85.

22. Atkinson EG, Dalvie S, Pichkar Y, Kalungi A, Majara L, Stevenson A, et al. Genetic structure correlates with ethnolinguistic diversity in eastern and southern Africa. Am J Hum Genet. 2022 Sep 1;109(9):1667–79.

23. Gurdasani D, Carstensen T, Tekola-Ayele F, Pagani L, Tachmazidou I, Hatzikotoulas K, et al. The African Genome Variation Project shapes medical genetics in Africa. Nature. 2015 Jan;517(7534):327–32.

24. Llorente MG, Jones ER, Eriksson A, Siska V, Arthur KW, Arthur JW, et al. Ancient Ethiopian genome reveals extensive Eurasian admixture in Eastern Africa. Science. 2015 Nov 13;350(6262):820–2.

25. Pais LS, Snow H, Weisburd B, Zhang S, Baxter SM, DiTroia S, et al. seqr: A web-based analysis and collaboration tool for rare disease genomics. Hum Mutat. 2022 Mar 9;

26. Amberger JS, Bocchini CA, Schiettecatte F, Scott AF, Hamosh A. OMIM.org: Online Mendelian Inheritance in Man (OMIM®), an online catalog of human genes and genetic disorders. Nucleic Acids Res. 2015 Jan 28;43(Database issue):D789–98.

27. Philippakis AA, Azzariti DR, Beltran S, Brookes AJ, Brownstein CA, Brudno M, et al. The Matchmaker Exchange: a platform for rare disease gene discovery. Hum Mutat. 2015;36(10):915–21.

28. Rodan LH, Spillmann RC, Kurata HT, Lamothe SM, Maghera J, Jamra RA, et al. Phenotypic expansion of CACNA1C-associated disorders to include isolated neurological manifestations. Genet Med Off J Am Coll Med Genet. 2021 Oct;23(10):1922–32.

29. Royer-Bertrand B, Jequier Gygax M, Cisarova K, Rosenfeld JA, Bassetti JA, Moldovan O, et al. De novo variants in CACNA1E found in patients with intellectual disability, developmental regression and social cognition deficit but no seizures. Mol Autism. 2021 Oct 26;12(1):69.

30. Schalk A, Cousin MA, Dsouza NR, Challman TD, Wain KE, Powis Z, et al. De novo coding variants in the AGO1 gene cause a neurodevelopmental disorder with intellectual disability. J Med Genet [Internet]. 2021 Dec 14 [cited 2022 Mar 7]; Available from: https://jmg-bmj-com.ezp-prod1.hul.harvard.edu/content/early/2021/12/14/jmedgenet-2021-107751

31. Muenke M, Adeyemo A, Kruszka P. An electronic atlas of human malformation syndromes in diverse populations. Genet Med. 2016 Nov;18(11):1085–7.

32. Samocha KE, Kosmicki JA, Karczewski KJ, O’Donnell-Luria AH, Pierce-Hoffman E, MacArthur DG, et al. Regional missense constraint improves variant deleteriousness prediction [Internet]. bioRxiv; 2017 [cited 2022 Nov 28]. p. 148353. Available from: https://www.biorxiv.org/content/10.1101/148353v1

33. Kircher M, Witten DM, Jain P, O’Roak BJ, Cooper GM, Shendure J. A general framework for estimating the relative pathogenicity of human genetic variants. Nat Genet. 2014 Mar;46(3):310–5.

34. Martin AR, Teferra S, Möller M, Hoal EG, Daly MJ. The critical needs and challenges for genetic architecture studies in Africa. Curr Opin Genet Dev. 2018 Dec;53:113–20.

35. Peterson RE, Kuchenbaecker K, Walters RK, Chen CY, Popejoy AB, Periyasamy S, et al. Genome-wide Association Studies in Ancestrally Diverse Populations: Opportunities, Methods, Pitfalls, and Recommendations. Cell. 2019 Oct 17;179(3):589–603.

36. Gudmundsson S, Singer-Berk M, Watts NA, Phu W, Goodrich JK, Solomonson M, et al. Variant interpretation using population databases: Lessons from gnomAD. Hum Mutat. 2021 Dec 2;

37. Karczewski KJ, Francioli LC, Tiao G, Cummings BB, Alföldi J, Wang Q, et al. The mutational constraint spectrum quantified from variation in 141,456 humans. Nature. 2020;581(7809):434–43.

38. Mathias RA, Taub MA, Gignoux CR, Fu W, Musharoff S, O’Connor TD, et al. A continuum of admixture in the Western Hemisphere revealed by the African Diaspora genome. Nat Commun. 2016 Oct 11;7:12522.

39. Zakharia F, Basu A, Absher D, Assimes TL, Go AS, Hlatky MA, et al. Characterizing the admixed African ancestry of African Americans. Genome Biol. 2009 Dec 22;10(12):R141.

40. Caswell-Jin JL, Gupta T, Hall E, Petrovchich IM, Mills MA, Kingham KE, et al. Racial/ethnic differences in multiple-gene sequencing results for hereditary cancer risk. Genet Med Off J Am Coll Med Genet. 2018 Feb;20(2):234–9.

41. Lek M, Karczewski KJ, Minikel EV, Samocha KE, Banks E, Fennell T, et al. Analysis of protein-coding genetic variation in 60,706 humans. Nature. 2016 Aug 18;536(7616):285– 91.

42. Manrai AK, Funke BH, Rehm HL, Olesen MS, Maron BA, Szolovits P, et al. Genetic Misdiagnoses and the Potential for Health Disparities. N Engl J Med. 2016 Aug 18;375(7):655–65.

43. Popejoy AB, Ritter DI, Crooks K, Currey E, Fullerton SM, Hindorff LA, et al. The clinical imperative for inclusivity: Race, ethnicity, and ancestry (REA) in genomics. Hum Mutat. 2018 Nov;39(11):1713–20.

44. Negoro T, Okura H, Matsuyama A. Induced Pluripotent Stem Cells: Global Research Trends. BioResearch Open Access. 2017 Jun 1;6(1):63–73.

45. Bank W. Kenya Gender and Poverty Assessment 2015-2016: Reflecting on a Decade of Progress and the Road Ahead. World Bank; 2018.

46. Richards S, Aziz N, Bale S, Bick D, Das S, Gastier-Foster J, et al. Standards and guidelines for the interpretation of sequence variants: a joint consensus recommendation of the American College of Medical Genetics and Genomics and the Association for Molecular Pathology. Genet Med. 2015 May 1;17(5):405–24.

